# Anti-SARS-CoV-2 cellular immunity in 571 vaccinees assessed using an interferon-γ release assay

**DOI:** 10.1101/2021.12.14.21267039

**Authors:** Yoshifumi Uwamino, Masatoshi Wakui, Yoko Yatabe, Terumichi Nakagawa, Akiko Sakai, Toshinobu Kurafuji, Ayako Shibata, Yukari Tomita, Masayo Noguchi, Akiko Tanabe, Tomoko Arai, Akemi Ohno, Hiromitsu Yokota, Shunsuke Uno, Wakako Yamasawa, Yasunori Sato, Mari Ikeda, Akihiko Yoshimura, Naoki Hasegawa, Hideyuki Saya, Mitsuru Murata

## Abstract

Generation of antigen-specific memory T cells has been analyzed only for few coronavirus disease 2019 (COVID-19) vaccinees, whereas antibody titers have been serologically measured for a large number of individuals. Here, we assessed the anti-severe acute respiratory syndrome coronavirus 2 (SARS-CoV-2) cellular immune response in a large cohort using interferon (IFN)-γ release assays (IGRAs) based on short-term whole blood culture. The study included 571 individuals who received the viral spike (S) protein-expressing BNT162b2 mRNA SARS-CoV-2 vaccine. Serum IgG titers against the receptor-binding domain (RBD) of S protein were measured. Samples of 28 vaccinees were subjected to flow cytometry analysis of T cells derived from short-term whole blood culture. IFN-γ production triggered by S antigens was observed in most individuals 8 weeks after receiving the second dose of the vaccine, indicating acquisition of T cell memory responses. The frequencies of activated T cell subsets were strongly correlated with IFN-γ levels, supporting the usability of our approach. S antigen-stimulated IFN-γ levels were weakly correlated with anti-RBD IgG titers and associated with pre-vaccination infection and adverse reactions after the second dose. Our approach revealed cellular immunity acquired after COVID-19 vaccination, providing insights regarding the effects and adverse reactions of vaccination.

## Introduction

Currently, mass vaccination is underway worldwide to overcome the coronavirus disease 2019 (COVID-19) pandemic caused by severe acute respiratory syndrome coronavirus 2 (SARS-CoV-2). The BNT162b2 mRNA SARS-CoV-2 vaccine, which leads to the expression of the viral spike (S) protein in vaccinee cells, is used most frequently [1, 2]. Previously, the antigen-specific memory T cells in only a small number of samples (less than 50) have been assessed, whereas antibody titers have been serologically measured for many vaccinee samples. Although ELISpot assays for detection of antigen-reactive interferon (IFN)-γ production and flow cytometry assays for detection of antigen-specific T cell activation are conventionally used for investigating cellular immunity, the number of samples that can be tested is limited because of the laborious procedures involved [3–7]. To better understand the immunological aspects of the efficacy of mass COVID-19 vaccination, T cell responses should be assessed along with antibody responses using a larger number of vaccinee samples.

Similar to serological assays used as laboratory tests for screening of immunity against *Mycobacterium tuberculosis* or cytomegalovirus infection, interferon (IFN)-γ release assays (IGRAs) based on short-term whole blood culture enable detection of antigen-specific Th1 type responses for a large number of samples [8, 9]. Because of short-term culture, the assay results reflect the memory, but not the naïve T cells, reactive to the antigens used. Therefore, these IGRAs are useful for detecting the acquisition of cellular immunity due to vaccination as well as due to natural infection. Recently, the QuantiFERON SARS-CoV-2 kit (Qiagen, Hilden, Germany) and the SARS-CoV-2 IGRA kit (Euroimmun Medizinische Labordiagnostika, Luebeck, Germany) have been developed for IGRAs based on short-term whole blood culture to functionally detect anti-SARS-CoV-2 T cells [10–13]. A small feasibility study involving 12 vaccinated individuals has demonstrated the ability of the QuantiFERON SARS-CoV-2 assay to identify T cell responses in COVID-19 vaccinees [10]. Two other studies evaluated the performance of the QuantiFERON SARS-CoV-2 assay in 18 and 22 vaccinated health care workers using samples collected only after vaccination [14, 15]. A recent study assessed cellular immunity acquired by 32 healthy individuals, 58 patients on hemodialysis, 29 patients on peritoneal dialysis, and 90 renal allograft recipients who had received the mRNA vaccine using the QuantiFERON SARS-CoV-2 assay [16]. Impairment of immune responses in 106 and 543 vaccinated patients undergoing hemodialysis was assessed using the QuantiFERON SARS-CoV-2 assay in two independent studies [17, 18]. Although the findings from these studies are interesting, they should be supported by observations obtained after using more samples from healthy individuals or by comparing the results obtained using the QuantiFERON SARS-CoV-2 assay with those of conventional assays (ELISpot and flow cytometry).

Understanding of not only humoral but also cellular immunity acquired after vaccination is expected to provide insights regarding common or distinct features of immunity in COVID-19 and other viral diseases as well as the effects of immunization. The present study aimed to assess cellular immunity in a large number of individuals who had received the BNT162b2 mRNA SARS-CoV-2 vaccine using IGRAs based on short-term whole blood culture. To confirm the usability of the IGRA based on its correlation with conventional assays, some randomly selected vaccinee samples were simultaneously subjected to flow cytometric analyses of T cells derived from the culture for IGRAs.

## Results

Of the 593 participants selected in the study, 10 participants who did not receive both doses of the vaccine and 10 participants from whom post-vaccination samples were not collected were excluded from further analyses. The remaining vaccinees were analyzed, except for two vaccinees for whom the assay did not work. The median age of the 571 analyzed vaccinees was 45 years [interquartile range (IQR), 36–53 years]. The vaccinees included 169 males (29.6%) and 402 females (70.4%). Eight individuals had a history of COVID-19 prior to vaccination (Table 1).

**Table 1.**
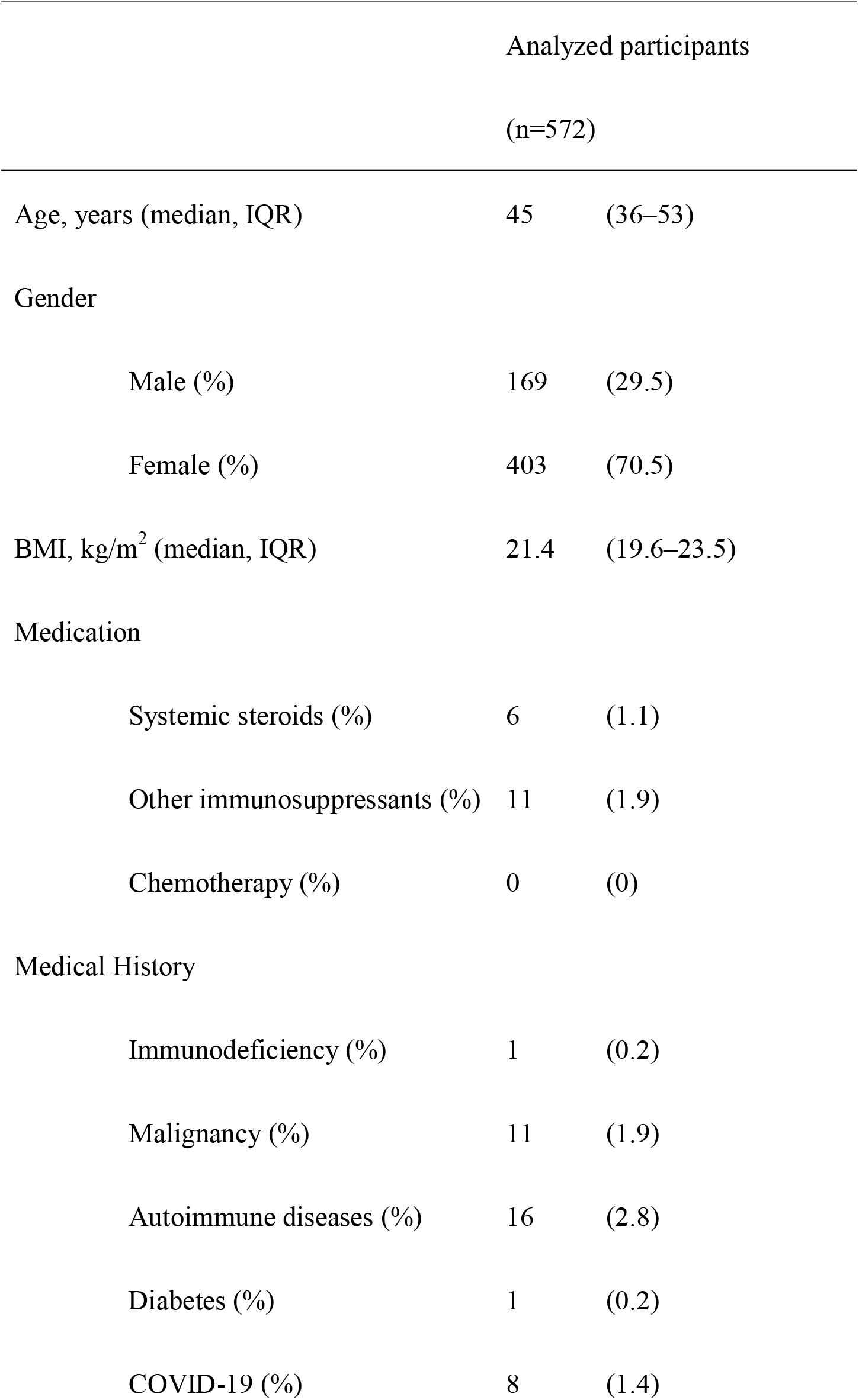
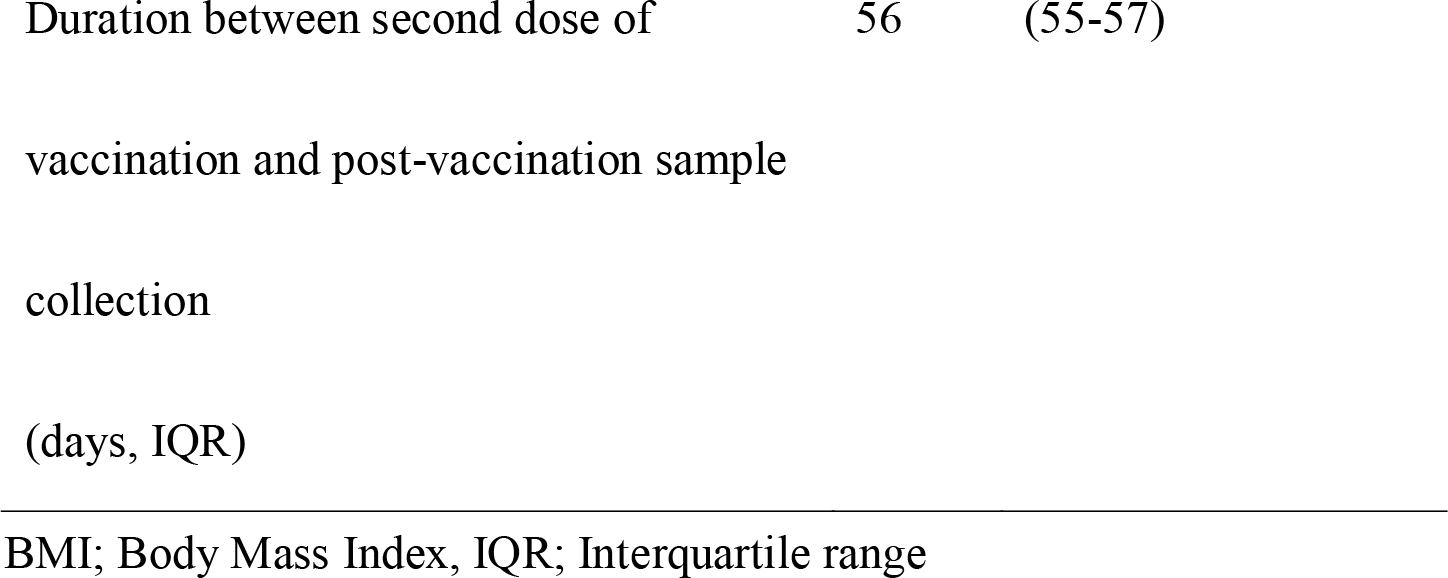
Demography of the analyzed participants.

### IGRA based on short-term whole blood culture (QuantiFERON SARS-CoV-2 assay)

To measure T cell responses, blood samples were stimulated with two peptide antigen pools. Antigen 1 (Ag 1) was designed to present epitopes present in the S1 subunit, which are recognized mainly by CD4^+^ T cells. Antigen 2 (Ag 2) was designed to present epitopes present in S1 and S2 subunits, which are recognized by CD4^+^ T cells and CD8^+^ T cells, respectively. In post-vaccine samples, the IFN-γ levels in the supernatant of short-term whole blood culture upon Ag 1 or Ag 2 stimulation differed widely (Fig. 1). The median IFN-γ levels in post-vaccine samples were higher than those in pre-vaccine samples upon both Ag 1 and Ag 2 stimulation. In case of Ag 1 stimulation, the pre-vaccination samples contained 0.00 IU/mL IFN-γ (IQR: −0.01 to 0.00 IU/mL), while the post-vaccination samples contained 0.35 IU/mL IFN-γ (IQR: 0.14–0.35 IU/mL) (*p* < 0.001). In case of Ag 2 stimulation, the pre-vaccination samples contained 0.00 IU/mL IFN-γ (IQR: −0.01 IU/mL to 0.01 IU/mL), while the post-vaccination samples contained 0.61 IU/mL IFN-γ (IQR: 0.26–1.32 IU/mL) (*p* < 0.001). These results were indicative of anti-SARS-CoV-2 Th1 memory responses acquired by vaccination.

**Figure 1.**
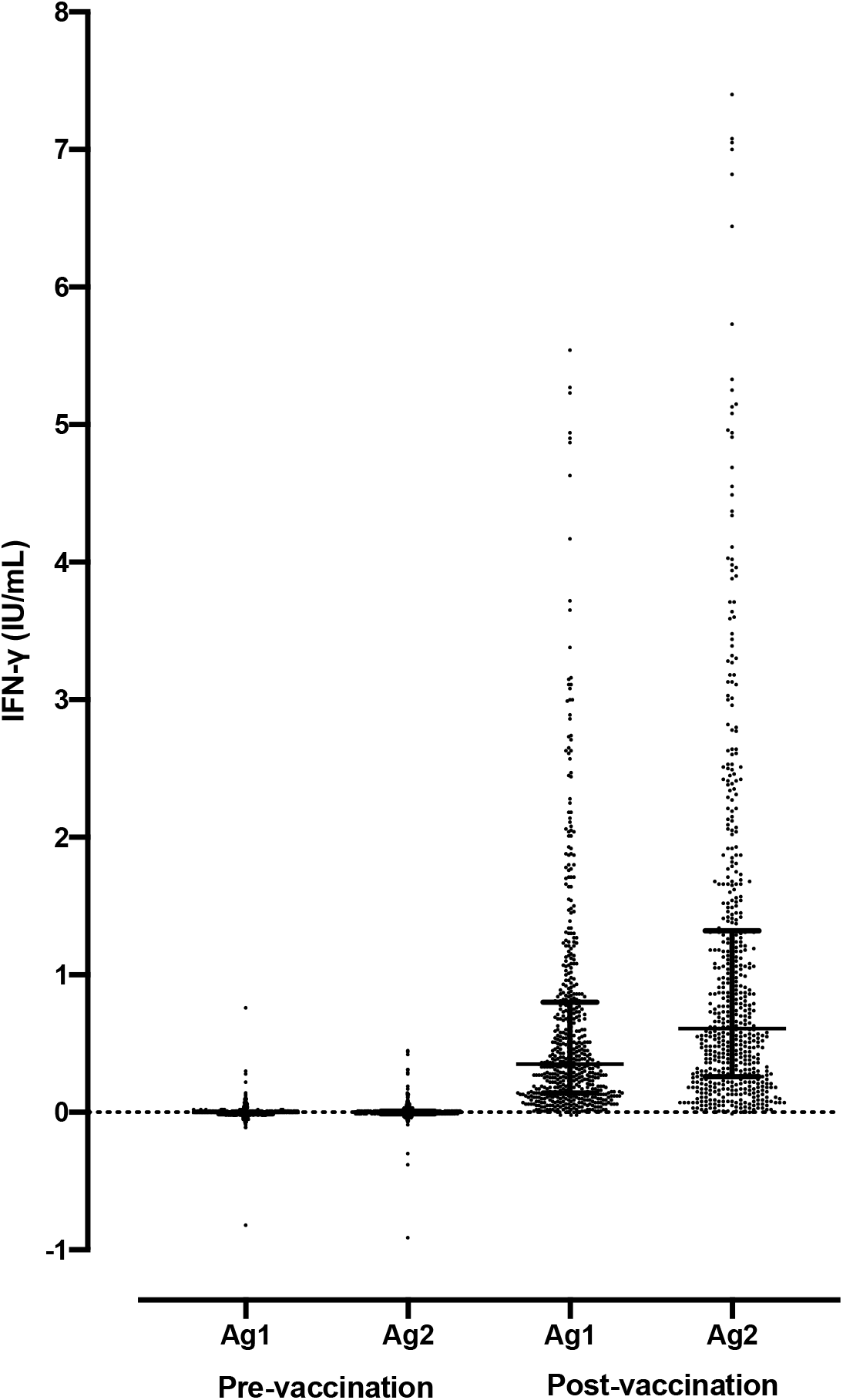
IFN-γ levels detected using QuantiFERON SARS-CoV-2 assays in pre- and post-vaccination samples. The distributions of IFN-γ levels induced by antigen 1 (Ag 1) or antigen 2 (Ag 2) stimulation are shown.

### Flow cytometry analyses for detecting anti-SARS-CoV-2 memory T cell responses

In 27 of the 28 randomly selected vaccinees, post-vaccination anti-SARS-CoV-2 memory T cell responses were determined using flow cytometry analyses. The percentages of CD3^+^ CD4^+^ CD134^+^ CD137^+^ cells, representing activated CD4^+^ T cell subsets, and the percentages of CD3^+^ CD8^+^ CD169^+^ CD137^+^ cells, representing activated CD8^+^ T cell subsets, increased in short-term whole blood culture after Ag 1 or Ag 2 stimulation for IGRAs (Fig. 2, Fig. E1). In addition, these percentages were strongly correlated with the IFN-γ levels in QuantiFERON SARS-CoV-2 assays (Fig. 3), indicating the usability of the QuantiFERON SARS-CoV-2 assay. A significant correlation of the percentages of activated T cells with the antibody titers or with the vaccinee age was not observed (Fig. E2).

**Figure 2.**
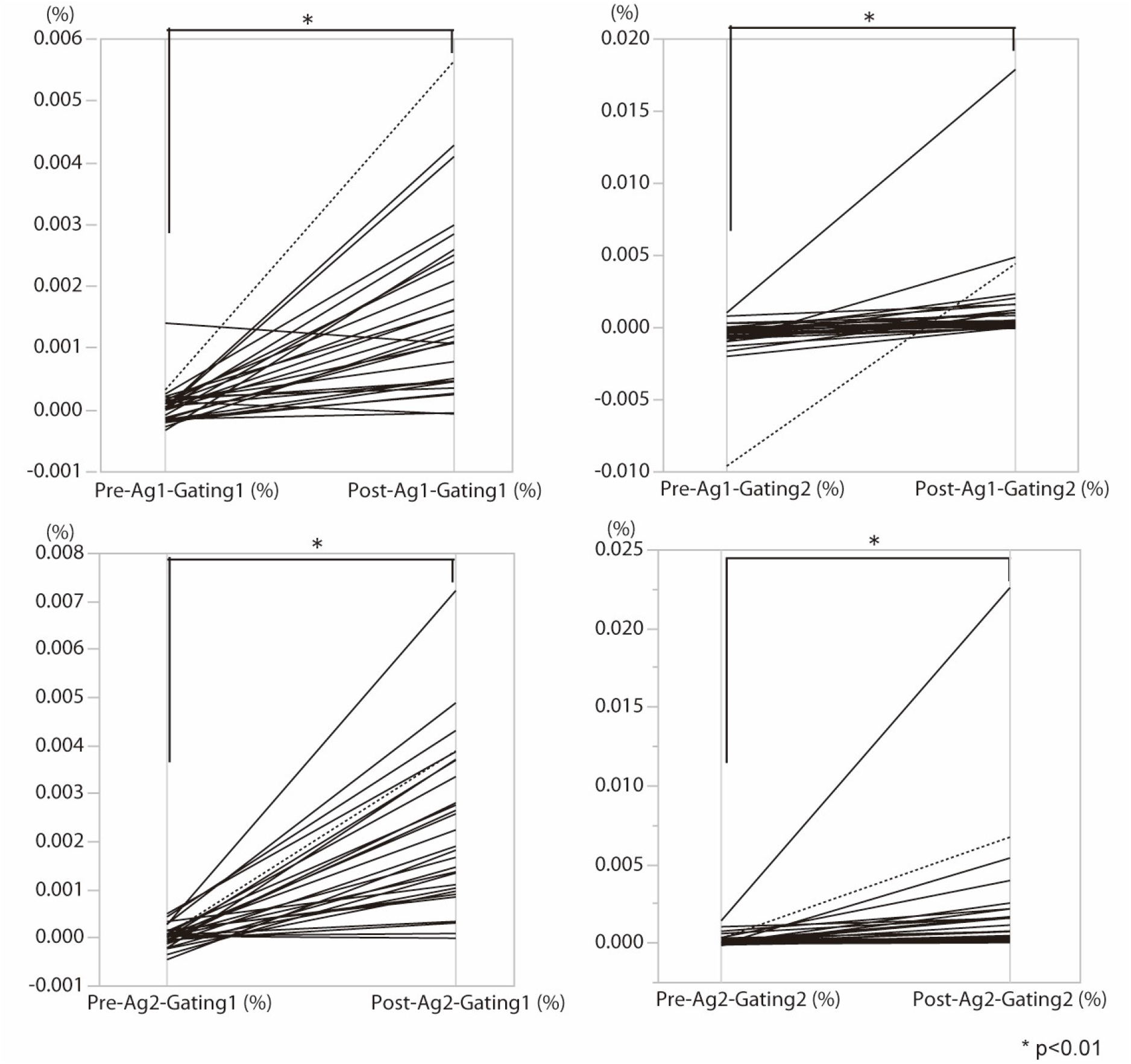
Distribution of frequencies of activated T cell subsets detected using flow cytometry analyses in pre- and post-vaccination samples. The distributions of frequencies of activated T cell subsets induced by antigen 1 (Ag 1) or antigen 2 (Ag 2) are shown.

**Figure 3.**
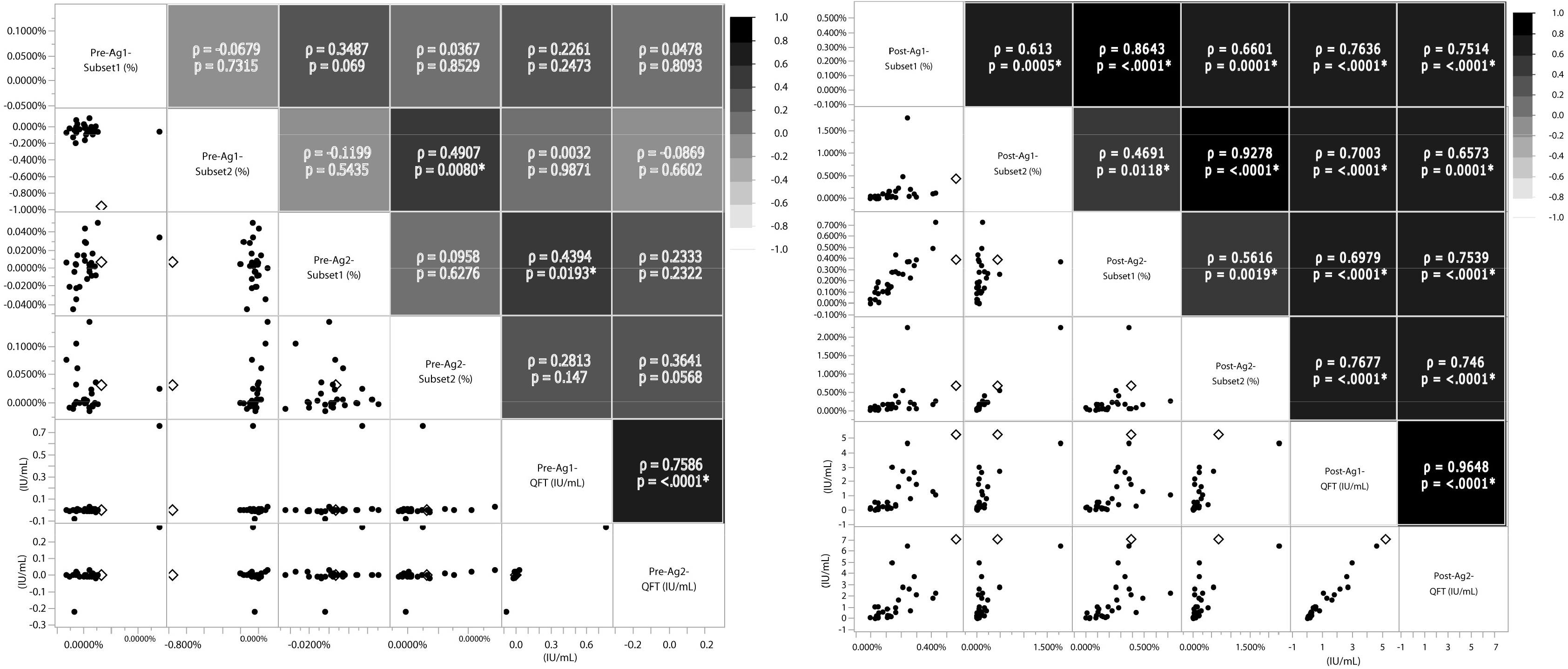
Correlation matrices for a set of flow cytometry and QuantiFERON SARS-CoV-2 data.

### Correlation of QuantiFERON SARS-CoV-2 data with antibody titers and epidemiological data

The IFN-γ levels in the supernatant of short-term whole blood culture after Ag 1 or Ag 2 stimulation demonstrated a weak positive correlation with antibody titers (IFN-γ levels upon Ag 1 stimulation versus antibody titers; r_s_ = 0.1863, *p* < 0.001; IFN-γ levels upon Ag 2 stimulation versus antibody titers; r_s_ = 0.1687, *p* < 0.001) (Fig. 4).

**Figure 4.**
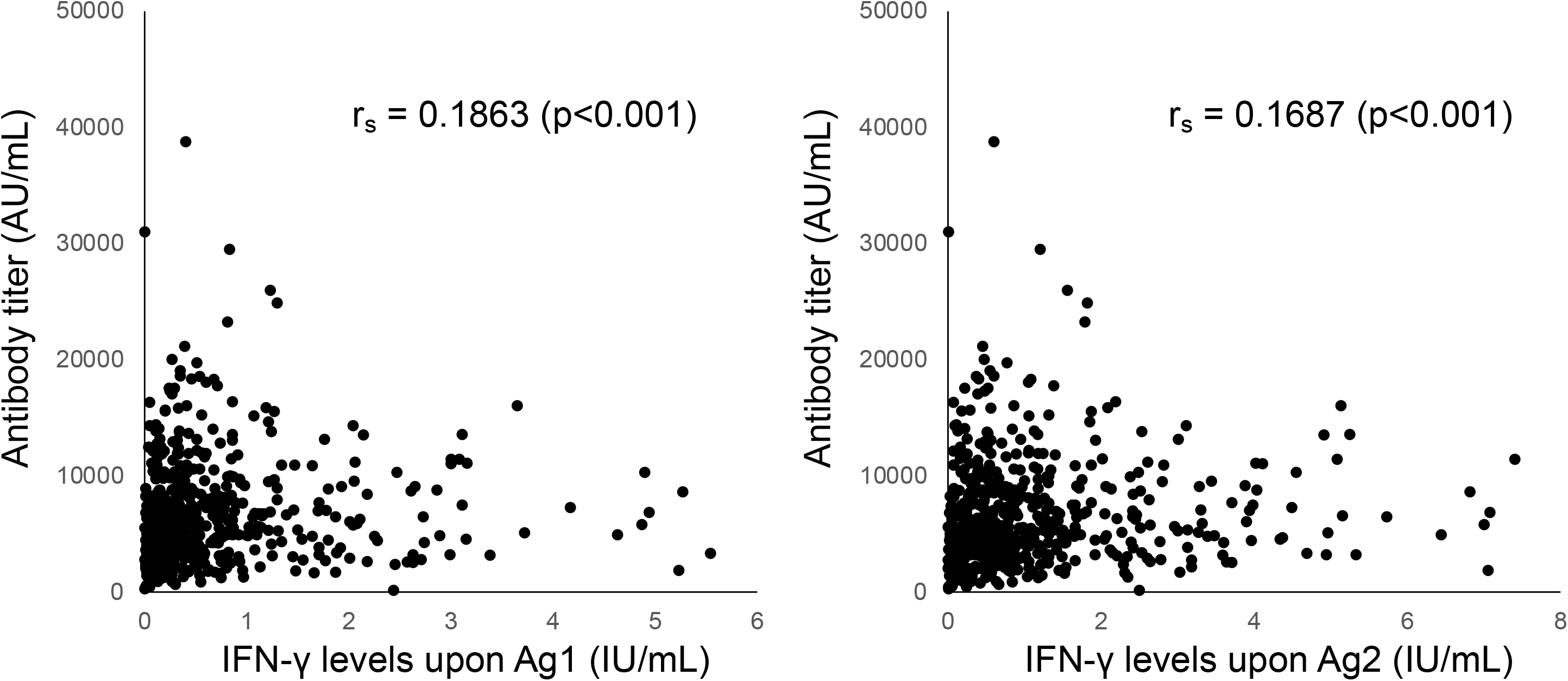
Correlations between IFN-γ levels induced by antigen 1 or antigen 2 stimulation versus antibody titers. The correlations between IFN-γ levels upon antigen 1 (Ag 1) or antigen 2 (Ag 2) stimulation versus anti-spike protein receptor-binding domain (RBD) IgG titers (antibody titers) are shown.

Individuals with a history of pre-vaccination COVID-19 presented higher IFN-γ levels resulting from Ag 1 stimulation than those without the history. IFN-γ level was 0.8 IU/mL (IQR 0.41–2.48 IU/mL) in individuals with a history of COVID-19 and 0.34 IU/mL IFN-γ (IQR 0.14–0.7925 IU/mL, *p* = 0.046) in those without a history of COVID-19. The IFN-γ levels resulting from Ag 2 stimulation also tended to be higher in individuals with history of the disease than in those without the history, although the difference was not significant (*p* = 0.051).

Vaccinees experiencing adverse reactions after the second vaccine dose (irrespective of symptom types) presented higher IFN-γ levels resulting from Ag 2 stimulation. The IFN-γ levels in vaccinees with adverse reactions after the second dose was 0.63 IU/mL (IQR, 0.27–1.42 IU/mL), while that in vaccinees without adverse reactions after the second dose was 0.53 IU/mL (IQR 0.19–1.11 IU/mL (*p* = 0.026). A similar tendency was observed with IFN-γ levels resulting from Ag 1 stimulation, albeit without statistical significance (*p* = 0.065). Fever, general fatigue, and local reactions were correlated with higher IFN-γ production upon Ag 1 and Ag 2 stimulation; these correlations were significant (Table 2).

**Table 2.**
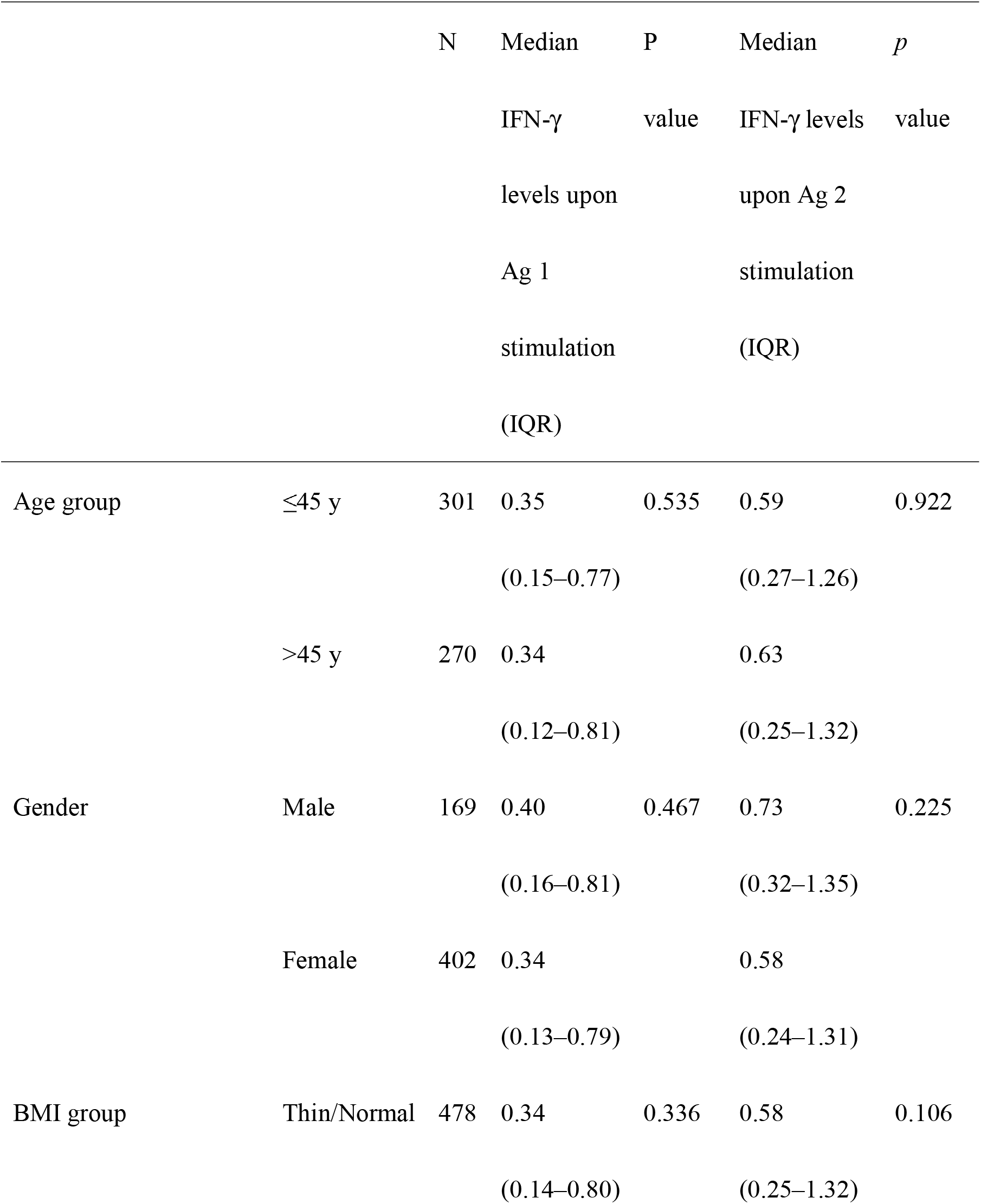
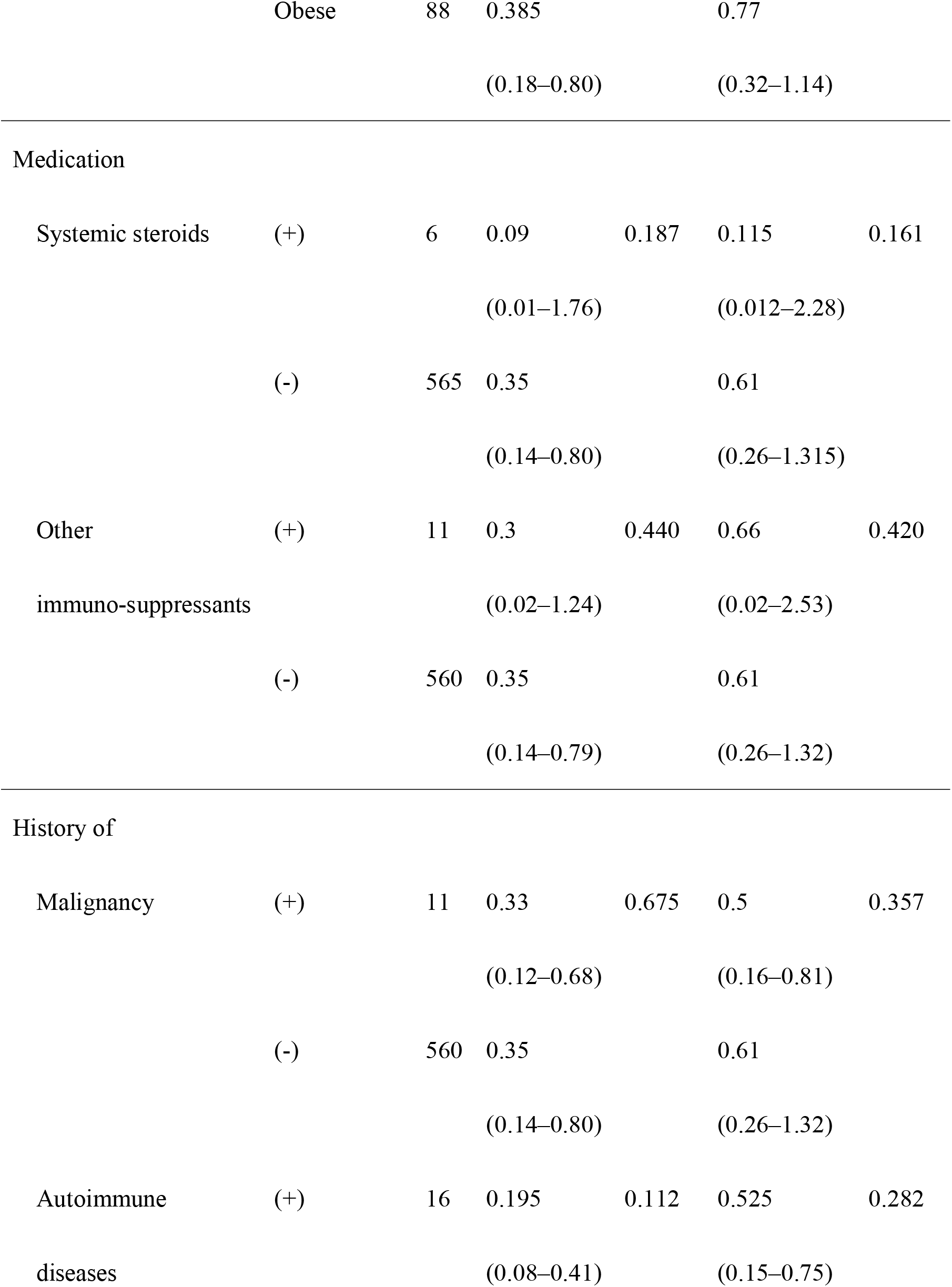
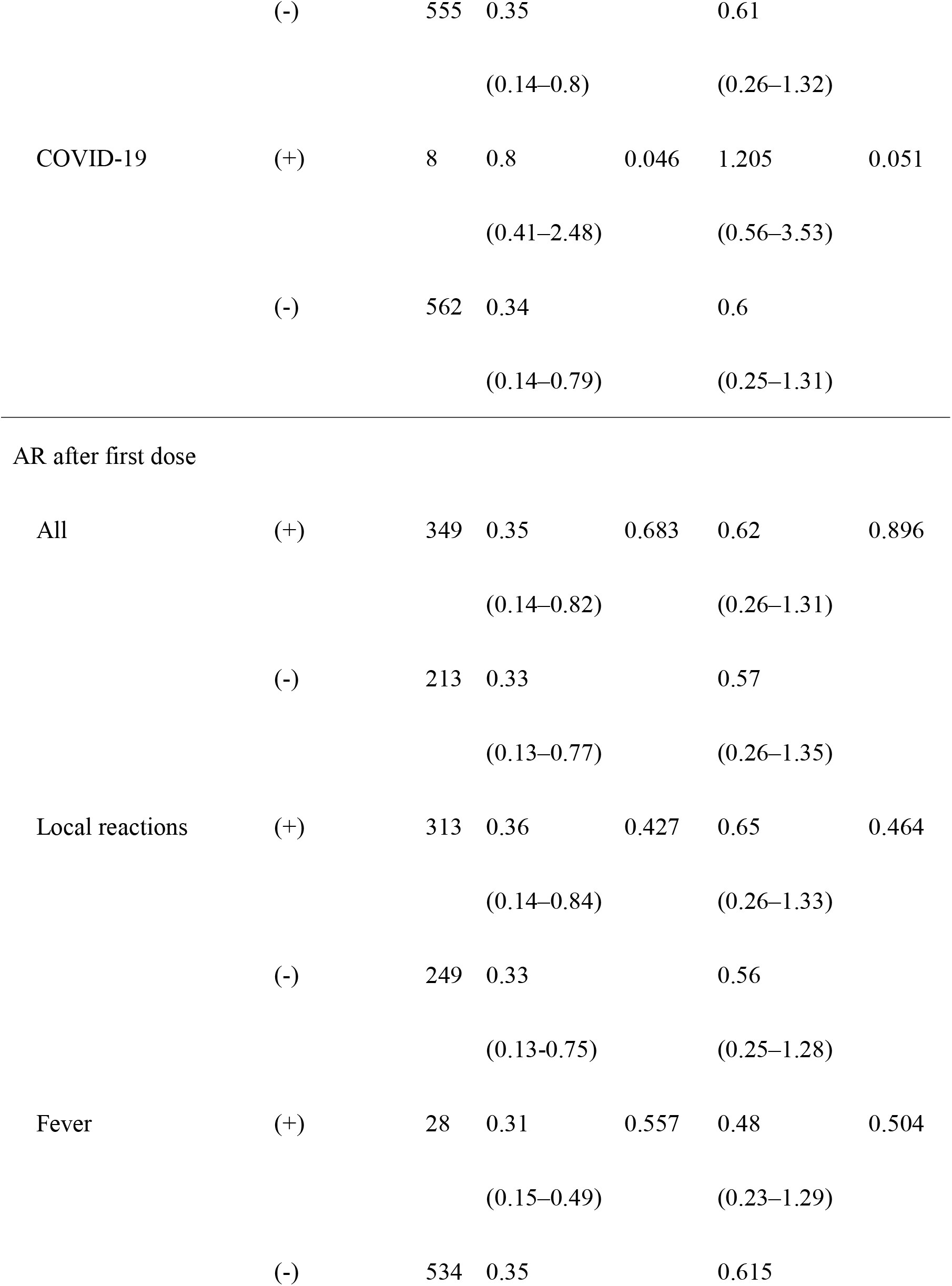
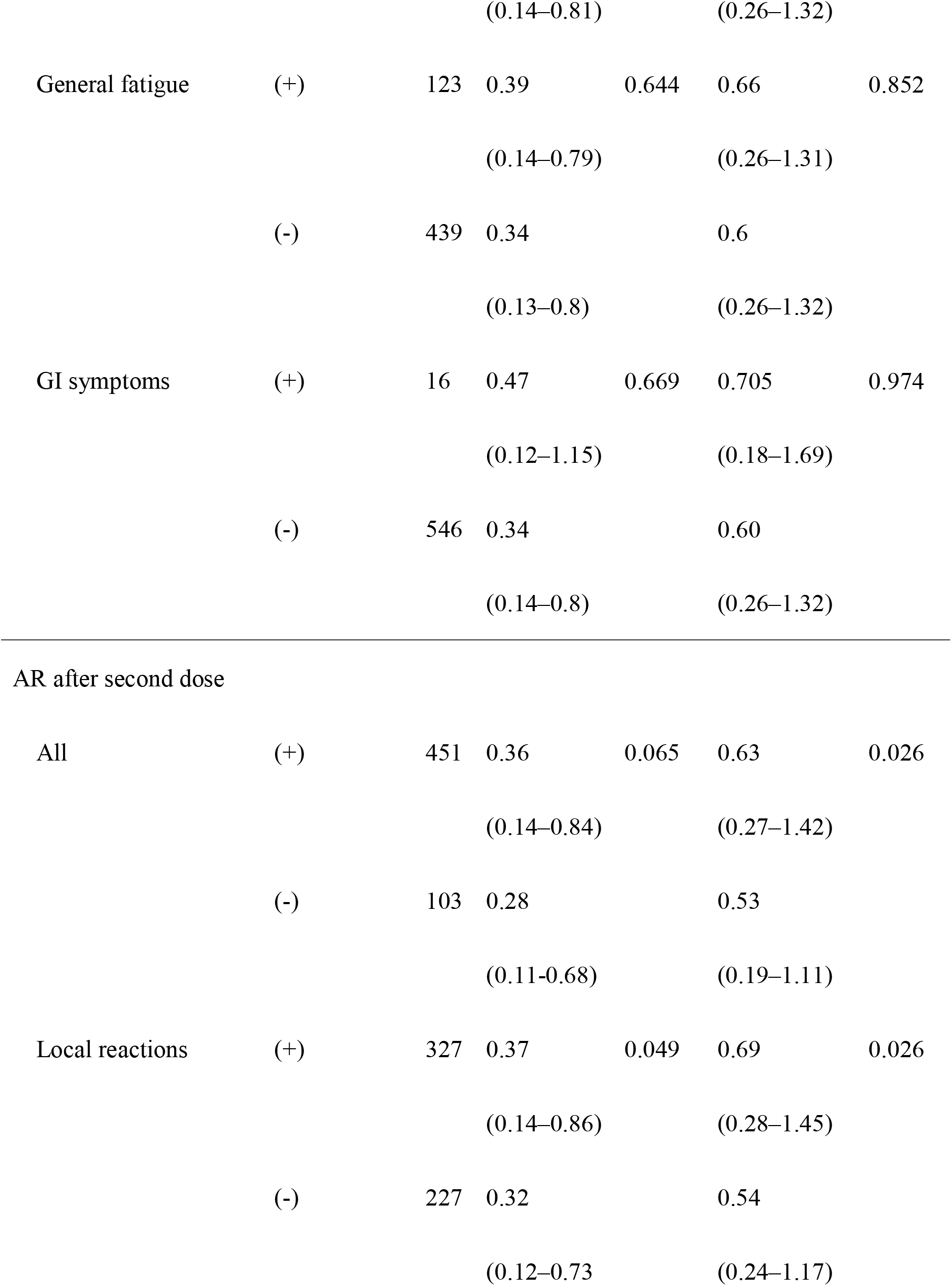
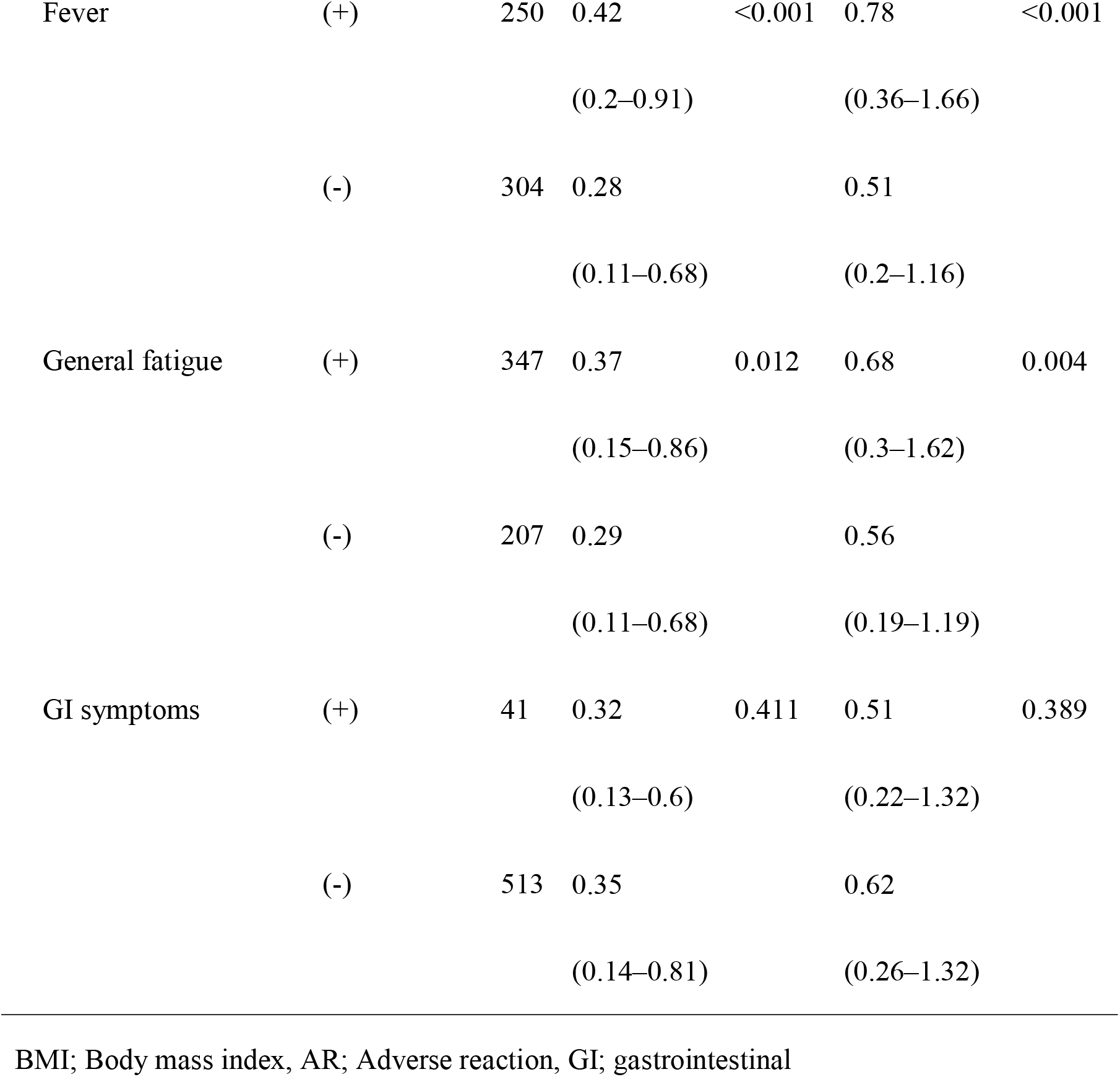
Differences in IFN-γ levels in post-vaccine samples between groups according to participants’ demographic parameters and adverse reactions.

| | | N | Median<br>IFN-γ<br>levels upon<br>Ag 1<br>stimulation<br>(IQR) | P<br>value | Median<br>IFN-γ levels<br>upon Ag 2<br>stimulation<br>(IQR) | *p*<br>value |
| --- | --- | --- | --- | --- | --- | --- |
| Age group | $\leq 45$ y | 301 | 0.35<br>(0.15–0.77) | 0.535 | 0.59<br>(0.27–1.26) | 0.922 |
| | $> 45$ y | 270 | 0.34<br>(0.12–0.81) | | 0.63<br>(0.25–1.32) | |
| Gender | Male | 169 | 0.40<br>(0.16–0.81) | 0.467 | 0.73<br>(0.32–1.35) | 0.225 |
|  | Female | 402 | 0.34<br>(0.13–0.79) |  | 0.58<br>(0.24–1.31) |  |
| BMI group | Thin/Normal | 478 | 0.34<br>(0.14–0.80) | 0.336 | 0.58<br>(0.25–1.32) | 0.106 |
|  | Obese | 88 | 0.385 |  | 0.77 |  |
|  |  |  | (0.18–0.80) |  | (0.32–1.14) |  |
| Medication |  |  |  |  |  |  |
| Systemic steroids | (+) | 6 | 0.09 | 0.187 | 0.115 | 0.161 |
|  |  |  | (0.01–1.76) |  | (0.012–2.28) |  |
|  | (-) | 565 | 0.35 |  | 0.61 |  |
|  |  |  | (0.14–0.80) |  | (0.26–1.315) |  |
| Other | (+) | 11 | 0.3 | 0.440 | 0.66 | 0.420 |
| immuno-suppressants |  |  | (0.02–1.24) |  | (0.02–2.53) |  |
|  | (-) | 560 | 0.35 |  | 0.61 |  |
|  |  |  | (0.14–0.79) |  | (0.26–1.32) |  |
| History of |  |  |  |  |  |  |
| Malignancy | (+) | 11 | 0.33 | 0.675 | 0.5 | 0.357 |
|  |  |  | (0.12–0.68) |  | (0.16–0.81) |  |
|  | (-) | 560 | 0.35 |  | 0.61 |  |
|  |  |  | (0.14–0.80) |  | (0.26–1.32) |  |
| Autoimmune | (+) | 16 | 0.195 | 0.112 | 0.525 | 0.282 |
| diseases |  |  | (0.08–0.41) |  | (0.15–0.75) |  |
|  | (-) | 555 | 0.35 |  | 0.61 |  |
|  |  |  | (0.14–0.8) |  | (0.26–1.32) |  |
| COVID-19 | (+) | 8 | 0.8 | 0.046 | 1.205 | 0.051 |
|  |  |  | (0.41–2.48) |  | (0.56–3.53) |  |
|  | (-) | 562 | 0.34 |  | 0.6 |  |
|  |  |  | (0.14–0.79) |  | (0.25–1.31) |  |
| AR after first dose |  |  |  |  |  |  |
| All | (+) | 349 | 0.35 | 0.683 | 0.62 | 0.896 |
|  |  |  | (0.14–0.82) |  | (0.26–1.31) |  |
|  | (-) | 213 | 0.33 |  | 0.57 |  |
|  |  |  | (0.13–0.77) |  | (0.26–1.35) |  |
| Local reactions | (+) | 313 | 0.36 | 0.427 | 0.65 | 0.464 |
|  |  |  | (0.14–0.84) |  | (0.26–1.33) |  |
|  | (-) | 249 | 0.33 |  | 0.56 |  |
|  |  |  | (0.13–0.75) |  | (0.25–1.28) |  |
| Fever | (+) | 28 | 0.31 | 0.557 | 0.48 | 0.504 |
|  |  |  | (0.15–0.49) |  | (0.23–1.29) |  |
|  | (-) | 534 | 0.35 |  | 0.615 |  |
|  |  |  | (0.14–0.81) |  | (0.26–1.32) |  |
| General fatigue | (+) | 123 | 0.39 | 0.644 | 0.66 | 0.852 |
|  |  |  | (0.14–0.79) |  | (0.26–1.31) |  |
|  | (-) | 439 | 0.34 |  | 0.6 |  |
|  |  |  | (0.13–0.8) |  | (0.26–1.32) |  |
| GI symptoms | (+) | 16 | 0.47 | 0.669 | 0.705 | 0.974 |
|  |  |  | (0.12–1.15) |  | (0.18–1.69) |  |
|  | (-) | 546 | 0.34 |  | 0.60 |  |
|  |  |  | (0.14–0.8) |  | (0.26–1.32) |  |
| AR after second dose |  |  |  |  |  |  |
| All | (+) | 451 | 0.36 | 0.065 | 0.63 | 0.026 |
|  |  |  | (0.14–0.84) |  | (0.27–1.42) |  |
|  | (-) | 103 | 0.28 |  | 0.53 |  |
|  |  |  | (0.11–0.68) |  | (0.19–1.11) |  |
| Local reactions | (+) | 327 | 0.37 | 0.049 | 0.69 | 0.026 |
|  |  |  | (0.14–0.86) |  | (0.28–1.45) |  |
|  | (-) | 227 | 0.32 |  | 0.54 |  |
|  |  |  | (0.12–0.73) |  | (0.24–1.17) |  |
| Fever | (+) | 250 | 0.42 | <0.001 | 0.78 | <0.001 |
|  |  |  | (0.2–0.91) |  | (0.36–1.66) |  |
|  | (-) | 304 | 0.28 |  | 0.51 |  |
|  |  |  | (0.11–0.68) |  | (0.2–1.16) |  |
| General fatigue | (+) | 347 | 0.37 | 0.012 | 0.68 | 0.004 |
|  |  |  | (0.15–0.86) |  | (0.3–1.62) |  |
|  | (-) | 207 | 0.29 |  | 0.56 |  |
|  |  |  | (0.11–0.68) |  | (0.19–1.19) |  |
| GI symptoms | (+) | 41 | 0.32 | 0.411 | 0.51 | 0.389 |
|  |  |  | (0.13–0.6) |  | (0.22–1.32) |  |
|  | (-) | 513 | 0.35 |  | 0.62 |  |
|  |  |  | (0.14–0.81) |  | (0.26–1.32) |  |

BMI; Body mass index, AR; Adverse reaction, GI; gastrointestinal

The ANCOVA model demonstrated that history of vaccination before developing COVID-19 and adverse reactions after the second dose were independently related to higher IFN-γ production upon Ag 1 and Ag 2 stimulation. In contrast, age, sex, obesity, or adverse reactions after the first dose were not correlated with higher IFN-γ levels (Table 3).

**Table 3.**
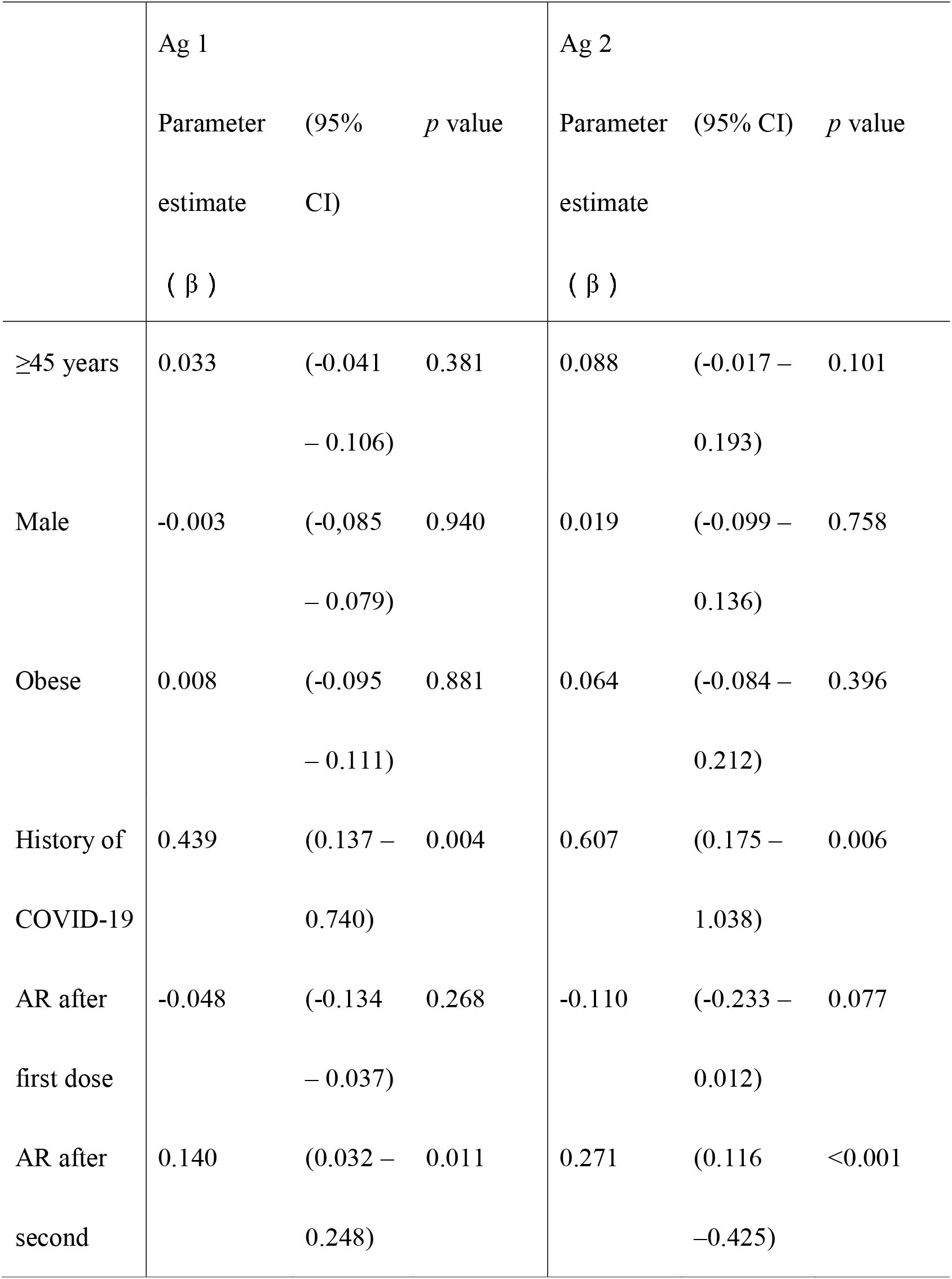
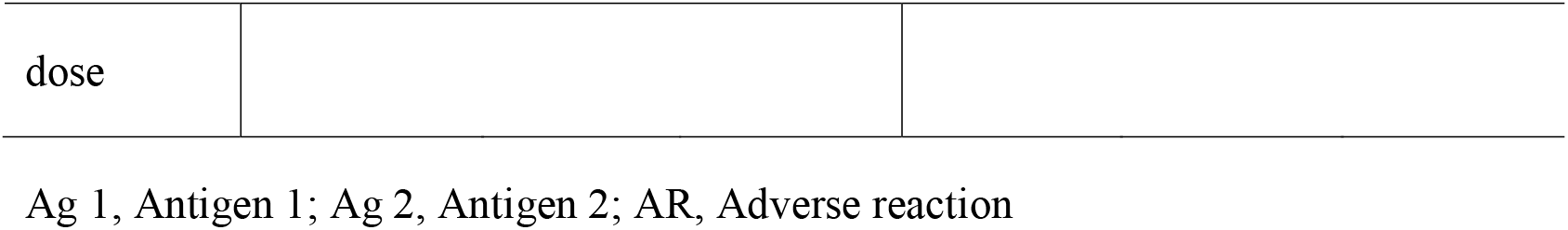
Relation between participants’ demographic data, adverse reactions, and IFN-γ levels.

## Discussion

IGRAs based on short-term whole blood culture, such as QuantiFERON SARS-CoV-2, can be used to measure the extent of helper T cell (Th)1 responsiveness to memory T cells and evaluate cellular immunity. The advantage of the test is that unlike ELISpot and flow cytometry, a large number of samples can be handled simultaneously. To our knowledge, this study is the first to demonstrate a correlation between QuantiFERON SARS-CoV-2 assay and flow cytometry data, supporting the usability of such IGRAs. Our results obtained using a large number of vaccinee samples were consistent with the results of previous studies obtained using small number of samples, proving that QuantiFERON SARS-CoV-2 can be used for assessing herd immunity [3–7].

Humoral responses depend on Th responses, which include not only Th1 but also other Th responses, such as those of Th2 and follicular helper T (Tfh) cells. Antigen-specific IFN-γ production mainly represents Th1 responses. Different Th responses do not exert synergistic effects on antibody production [19]. Rather, Th2 responses antagonize Th1 responses. Thus, the contribution of other Th responses to humoral responses might result in significant but weak correlations between the IgG titers and IFN-γ levels. Recently, a multicenter observational study using the anti-RBD IgG titer assay and the QuantiFERON SARS-CoV-2 assay, as used in the present study, involving 543 patients on hemodialysis and 75 healthy volunteers, has been reported by Van Praet et al. [18]. In that study, the Spearman correlation coefficients between antibody titers and IFN-γ levels in patients on hemodialysis and in healthy volunteers were 0.57 and 0.44, respectively, which were stronger than the correlations observed in the present study. The large difference in sample numbers between our study and that of Van Praet et al. might have contributed to the discrepancy in results obtained with the healthy volunteers, while some differences in the quality of immune responses of the 571 individuals of this study and that of the 543 patients on hemodialysis reported by Van Praet et al. might have resulted in the discrepancy in correlations.

Observations from a small number of vaccinees who had developed COVID-19 prior to receiving vaccination in the present study indicated that vaccination after SARS-CoV-2 infection enhanced acquisition of cellular immunity, as has been reported previously regarding humoral immunity [20]. Thus, vaccination possibly boosted cellular immunity acquired by past infection. To assess whether and how anti-SARS-CoV-2 memory T cells are maintained in vivo, further studies using QuantiFERON SARS-CoV-2 and individuals vaccinated before contracting COVID-19 are required.

Studies have demonstrated an association between antibody titers and adverse reactions, especially after the second vaccine dose, although the results of other studies do not support this observation [21, 22]. In the present study, the antigen-specific IFN-γ production levels were associated with adverse reactions after the second dose but not after the first dose. While the absence of adverse reactions should not be believed to predict failure to acquire immunity in vaccinees, it is plausible that humoral and cellular responses acquired by the first vaccination contributed to the occurrence of some adverse reactions following the second vaccination.

Currently, the necessity of additional vaccination is being decided mainly based on antibody titers. However, not only antibodies, but also memory B and T cells, participate in immune memory responses. In a previous report, the cut-off value of QuantiFERON SARS-CoV-2 was determined using results of healthy controls before (considered negative) and after vaccination (considered positive) [16]. While this cut-off value may assist in distinguishing between the absence and presence of cellular immunity, it cannot be used for assessing whether vaccination is sufficient for preventing SARS-CoV-2 infection. From the practical perspective, determination of the IFN-γ level in IGRAs for determining whether additional vaccination is required is critical for better infection control. To address this concern, correlations between IFN-γ levels and occurrence of breakthrough infection and comparison of IFN-γ levels between patients with severe and non-severe disease post-vaccination might be helpful.

Ag 1 is a mixture of 13-mer peptides restricted to HLA class II, which activates CD4^+^, but not CD8^+^, T cells. In contrast, Ag 2 is a mixture of 8- and 13-mer peptides restricted to HLA class I and class II, respectively, that activates both CD8^+^ and CD4^+^ T cells. In the present study, the frequency of activated CD8^+^ T cells increased even upon Ag 1 stimulation, while the IFN-γ levels induced upon Ag 2 stimulation were expectedly higher than those induced upon Ag 1 stimulation. This suggests that cross-presentation of some peptides derived from Ag 1 by antigen-presenting cells might occur unexpectedly, resulting in activation of CD8^+^ T cells. There are also two possibilities of alternative manners other than classical cross-presentation. One is that some 13-mer peptides may be presented by direct binding to HLA class I despite being longer than the suitable length [23]. Another possibility is that non-canonical antigen trimming by intracellular aminopeptidases may occur to activate CD8+ T cells in the HLA class I-restricted manner [24]. Thus, it is plausible that IFN-γ release upon Ag 1 stimulation is caused not only by CD4^+^ T cells but also by CD8^+^ T cells. Although CD4^+^ T cell responses in patients on hemodialysis and healthy volunteers undergoing vaccination were assessed using Ag 1 stimulation in a recent study, the present study demonstrated that distinction between CD4^+^ and CD8^+^ T cell responses using Ag 1 and Ag 2 is challenging [17]. This observation cannot be obtained from studies using IGRA alone. Interestingly, flow cytometry analyses in the present study revealed potential cross-presentation of peptides designed to activate CD4^+^ T cells to CD8^+^ T cells, which occurs during short-term whole blood culture (Fig. 2, Fig. E1).

We noticed that a strong T cell responses with Ag1 and Ag2 were observed part of vaccinees with low antibody titer (Fig.4). This consists with reports showing a strong prevention of severe disease by vaccination by a robust T cell immunity even if antibody response was low [25, 26].

In conclusion, IGRAs based on short-term whole blood culture revealed cellular immunity acquired by COVID-19 vaccination in a large number of individuals, providing insights regarding the effects and adverse reactions of vaccination. Our findings showed that IGRAs based on short-term whole blood culture can be used for assessing herd immunity against SARS-CoV-2.

## Materials and methods

### Subjects and samples

This is a part of a long-term observational study of vaccinated Japanese cohorts. From February 16th to March 9th, 2021, staffs of Keio University School of Medicine and Keio University Hospital (Tokyo, Japan) were recruited and included in the study after obtaining written informed consent from all 673 participants who were willing to receive the vaccine before the start of mass vaccination. The study design was approved by the Ethics Committee of Keio University School of Medicine (20200330). The mass vaccination using the BNT162b2 vaccine (COMIRNATY intramuscular injection, Pfizer, New York, USA) started on March 5th, 2021. The second vaccine dose was administered 3 weeks after the first dose.

For this study, two serial samples collected from 593 participants selected according to inclusion criteria were used. Pre-vaccination samples were collected before or on the same day of receiving the first dose of the vaccine. The post-vaccination samples were collected at approximately 8 weeks after receiving the second dose.

### Collection of epidemiological data

Before collecting pre-vaccination samples, a questionnaire was provided to all participants and information regarding age, sex, height, body weight, use of systemic steroids or other immunosuppressants, history of undergoing cancer chemotherapy, and history of immunodeficiency, malignancy, autoimmune diseases, diabetes, and COVID-19 was obtained. During collection of post-vaccination samples, adverse reactions after each vaccine dose were checked. Regarding adverse reactions, the vaccinees were enquired about the development of fever, general fatigue, local reactions, and gastrointestinal symptoms. The date of vaccination was obtained from the description of BNT162b2 vaccination in the vaccinees’ medical records.

### Determination of antibody titer

Immediately after sample collection, serum IgG titers against SARS-CoV-2 spike (S) protein S1 subunit receptor-binding domain (RBD) were measured using Alinity SARS-CoV-2 IgG II Quant reagents (Abbott Laboratories, Illinois, USA) and Alinity i Analyzer i (Abbott Laboratories, Illinois, USA) according to the manufacturer’s instructions.

### IGRA based on short-term whole blood culture

The QuantiFERON SARS-CoV-2 RUO assay kit (QFN) (Qiagen, Hilden, Germany) was used for the IGRA based on short-term whole blood culture. In this assay, two types of SARS-CoV-2 S peptides were pooled to stimulate T cells as antigens, namely Ag 1 and Ag 2. Whole blood samples collected in lithium heparin tubes were transferred to four QuantiFERON tubes coated with Ag 1, Ag 2, and mitogen (phytohemagglutinin) as positive controls and non-peptide substances as negative controls, respectively. After incubation at 37°C for 16−24 h, the cultured whole blood samples were processed according to the protocol mentioned by the manufacturer. In brief, the supernatants were collected, except for the samples from 28 vaccinees, samples from whom were subjected to flow cytometry analysis. The cultured whole blood samples from these 28 randomly selected vaccinees were transferred to other tubes to separate and collect both cells and supernatants via centrifugation.

The supernatants were subjected to ELISAs for measuring IFN-γ concentrations, which were performed using the AP-96 auto microplate EIA reader (Kyowa Medex, Tokyo, Japan) under optimal conditions for the QuantiFERON SARS-CoV-2 ELISA kit (Qiagen, Hilden, Germany).

### Flow cytometry assays for detecting T cell activation

Cells separated from the short-term whole blood culture of 28 randomly selected vaccinees were subjected to flow cytometry assays for detecting T cell activation. After immunostaining with fluorochrome-conjugated antibodies, hemolysis with ammonium chloride, and washing with buffers, the cells were analyzed in a BD FACSLyric cytometer (Becton and Dickinson, CA, USA). The fluorochrome-conjugated antibodies used for immunostaining included CD3 (LEU-4) FITC, CD4 APC-H7, CD8 PerCP-Cy5.5, CD69 PE, CD134 PE-Cy7, and CD137 APC (BD Biosciences, CA, USA). Flow cytometry data were analyzed using the BD FACSuite software (Becton and Dickinson). Activated CD4^+^ T cells were defined as CD3^+^ CD4^+^ CD134^+^ CD137^+^ cells, while activated CD8^+^ T cells were defined as CD3^+^ CD8^+^ CD169^+^ CD137^+^ cells [27, 28].

### Statistical analyses

The statistical data of the participants were generated using frequencies and proportions for categorical values and the median and IQR for continuous variables. Fisher’s exact test was used for categorical values and the Mann-Whitney U test was used for continuous variables. The IFN-γ levels of culture supernatants derived from Ag 1 and Ag 2 tubes were corrected by subtracting the values measured in negative controls. These corrected IFN-γ levels for Ag 1 or Ag2 were compared between pre- and post-vaccination samples. In addition, the correlation between IFN-γ levels and antibody titers was examined.

To identify factors affecting cellular immunity, IFN-γ levels for Ag 1 and Ag 2 after vaccination were compared according to patient characteristics, including sex, age (grouped into two groups according to median age of all the participants), body mass index (BMI) (grouped into two groups; BMI ≤ 25 kg/m^2^ as thin/normal and BMI > 25 kg/m^2^ as obese), medication (systemic steroids or other immunosuppressants), and history of malignancy, autoimmune diseases, and COVID-19. To investigate whether the occurrence of adverse reactions was related to immune responses acquired after vaccination, IFN-γ levels after vaccination were compared based on the episodes representing adverse reactions after each vaccine dose.

Finally, to identify factors independently associated with post-vaccination IFN-γ levels, a general linear model consisting of 6 categorical values, namely sex, age (more than 45 years versus 45 years or less), obesity, history of COVID-19, adverse reactions after the first dose, and adverse reactions after the second dose was built.

Regarding flow cytometric analysis, the percentages of CD3^+^ CD4^+^ CD134^+^ CD137^+^ phenotypic cells for Ag 1, Ag 2, and positive controls were corrected by subtracting the percentages for negative controls. The percentages of CD3^+^ CD8^+^ CD69^+^ CD137^+^ cells for Ag 1, Ag 2, and positive controls were corrected similarly. Data obtained from flow cytometric analysis and IGRA were compared using the Wilcoxon matched-pair signed-ranks test.

All the statistical analyses were performed using JMP version 15 (SAS Institute, North Carolina, USA). Two-sided *p*-values less than 0.05 were considered statistically significant.

## Data Availability

All data produced in the present study are available upon reasonable request to the authors.

## Abbreviations

COVID-19: coronavirus disease 2019
SARS-CoV-2: severe acute respiratory syndrome coronavirus 2
IFN: interferon
IGRA: IFN-γ release assay
RBD: receptor-binding domain

## Acknowledgments

This study was supported by the Research Funds of the Keio University School of Medicine and Grant from Public Foundation of the Vaccination Research Center, Japan (to Y.U. and M.W.). We thank the staff of Keio University Shinanomachi Campus for participating in the present study.

## Data availability statement

The data that support the findings of the present study are available from the corresponding author upon reasonable request.

## Ethics approval statement

The study design was approved by the Ethics Committee of Keio University School of Medicine (20200330).

## Patient consent

Written informed patient consent was obtained before the study.

## Author contributions

YU and MW contributed equally to this work, and both should be considered first authors. YU, MW, and MM conceived and designed the study. YU, MW, ASh, YT, AT, TA, AO, and WY recruited the participants. YY and MI optimized the protocol for the flow cytometric analysis. YY, ASa, TK, and MN assayed the samples and collected the data. YU, MW, YY, TN, YS, AY, and MM analyzed and interpreted the data. YU, MW, YY, and TN wrote the manuscript. YU, MW, YY, TN, ASa, TK, ASh, YT, MN, AT, TA, AO, HY, SU, WY, YS, MI, AY, NH, HS, and MM critically reviewed and revised the manuscript. All authors approved the final version of the manuscript to submit for publication.

