## Supplementary material for "Anti-SARS-CoV-2 cellular immunity in 571 vaccinees assessed using an interferon-γ release assay": Supplimental Figure legends

**Extended figure legends**

**Figure E1.** Representative data of the flow cytometric analysis for detecting activated T cells from the short-term whole blood culture. The data were obtained as described in the Materials and methods. The “CD4 3/69/8/134/137” staining was done for detecting activated CD4+ or CD8+ T cells from the culture with antigen peptides designed to stimulate CD4+ T cells (Ag 1). The “CD4+8 3/69/8/134/137” staining was done for detecting activated CD4+ or CD8+ T cells from the culture with both antigen peptides designed to stimulate CD4+ T cells and those designed to stimulate CD8+ T cells (Ag 2). The “negative 3/69/8/134/137” staining was done for detecting activated CD4+ or CD8+ T cells from the culture without antigen peptides as negative controls. The “positive 3/69/8/134/137” staining was done for detecting activated CD4+ or CD8+ T cells from the culture with mitogen (phytohemagglutinin) as positive controls.

**Figure E2.** Correlation matrices for a set of age, antibody titer, flow cytometry, and QuantiFERON SARS-CoV-2 data.
