## Supplementary figures and images for "Anti-SARS-CoV-2 cellular immunity in 571 vaccinees assessed using an interferon-γ release assay"

### Supplimental Figures

#### Figures E1 and E2

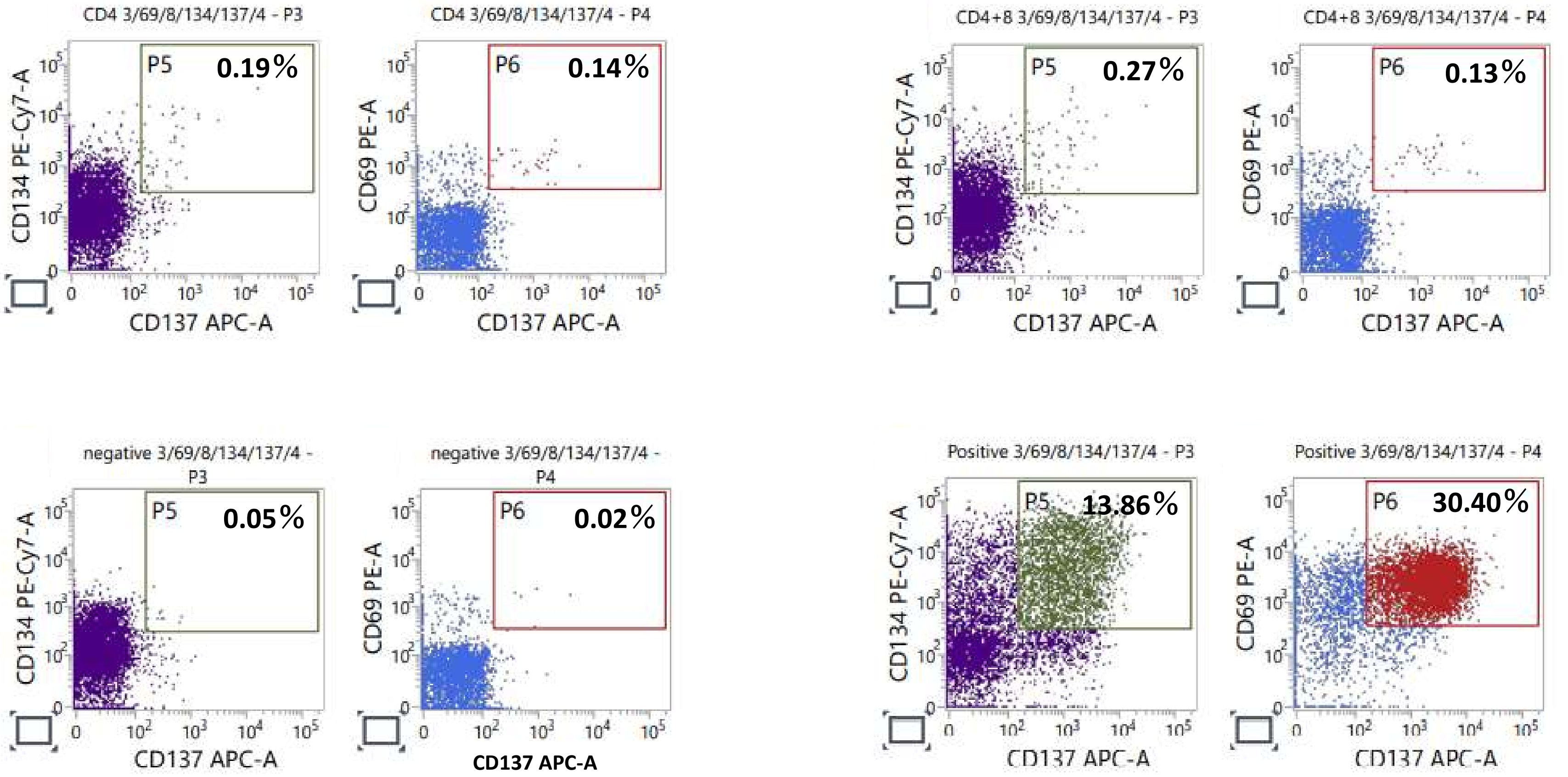

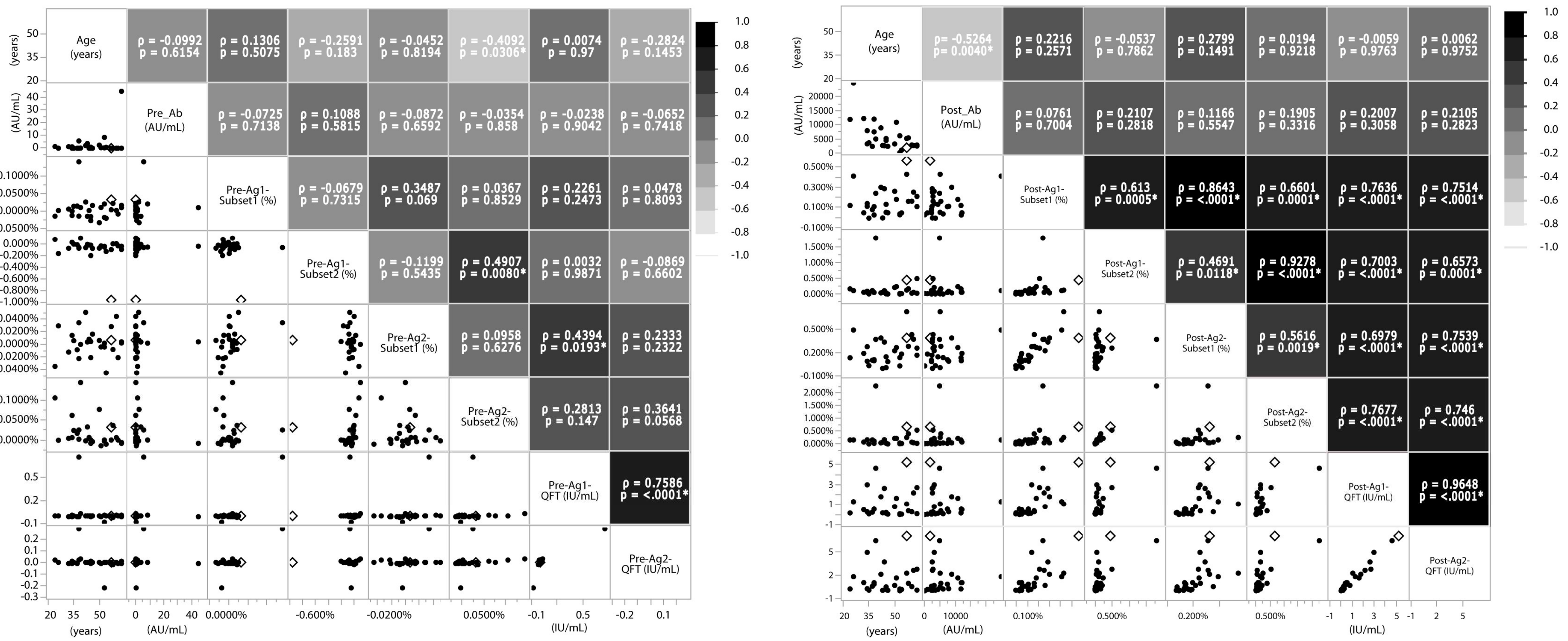
